# Factors associated with release relief of Long COVID symptoms at 12-Months and their impact on daily life

**DOI:** 10.1101/2022.11.18.22282459

**Authors:** Dominique Salmon, Dorsaf Slama, Françoise Linard, Nicolas Dumesges, Valérie Lebaut, Florence Hakim, Pauline Oustric, Emilie Seyrat, Patricia Thoreux, Esaie Marshall

## Abstract

**Introduction:** Our purpose was to describe the course of Long COVID symptoms after 12-month follow-up, their impact on daily life and the factors associated with the relief of symptoms.

**Methods:** A cross-sectional survey was conducted within an out-patient clinic for Long COVID patients. Participants, who had experienced their initial COVID-19 episode between January, 15 2020 and May 21, 2021, were contacted 12-months post onset. Their characteristics, symptom course at initial COVID-19 episode, Long COVID phase and one year follow-up along with remission status were collected through a questionnaire and a specific post COVID remission scale from complete remission to persistence of symptoms and dependence in daily life activities.

**Results:** Among the 231 long COVID participants who answered the 12-month follow-up questionnaire, 63.2% had developed SARS-CoV-2 antibodies before COVID-19 vaccination. At 12-month follow-up, only 8.7% of the participants felt in complete remission while 28.6% noted a significant improvement of their symptoms. The prevalence rate of most symptoms remained high at 12 months: asthenia 83.1%, neurocognitive and neurological symptoms 91.8%, cardiothoracic symptoms 77.9%, musculoskeletal 78.8%. During Long COVID phase, 62.2% had to stop working at least once and only 32.5% resumed professional activities full time at one year follow-up. The presence of SARS-CoV-2 antibodies before COVID-19 vaccination was associated with an increased probability of significant improvement at one year (aPRR: 1.60, p=0.028) while ageusia at initial Long COVID phase was associated with a lower probability of improvement (aPRR: 0.38, p=0.007).

**Conclusion:** While observing a trend towards some improvement in a majority of long COVID patients at a 12-month follow-up, fatigue, musculoskeletal pain, cardiothoracic symptoms and neurocognitive impairment persisted in most of them. Having developed SARS-CoV-2 antibodies was associated with a better prognosis while persistent ageusia at long COVID phase seems to be associated with the persistence of symptoms.

## Introduction

About 15% to 30% of the patients develop long-term symptoms after an acute symptomatic COVID-19 episode [1–3]. In October 2021, WHO defined this post COVID condition as long lasting symptoms including mainly fatigue, shortness of breath and palpitations, cognitive dysfunction but also damage to other organs [4]. These symptoms may be due to a new onset of the infection following initial recovery from an acute COVID-19 episode, or persist from the initial illness [4, 5, 6]. The lack of virological documentation of SARS-CoV-2 infection (PCR not made at the initial episode and/or negative serology) does not exclude this diagnosis [4]. These symptoms evolve in fluctuating waves and often get worse following physical or intellectual effort. This condition has been commonly called PACS (post-acute COVID syndrome) or Long COVID by the patients themselves.

Several factors associated with the occurrence of long-lasting symptoms have been identified (7, 8, 9). Unlike severe forms of COVID-19, which mostly affect men and the elderly, prolonged symptoms are more common in women [7, 8, 9] and relatively young subjects around the age of 45 [7]. Presenting a high number of symptoms during the initial COVID-19 episode appears to increase the risk of occurrence of long-term symptoms [7, 8]. The impact of SARS-CoV-2 antibody levels is controversial. According to varied studies, a high level of antibodies either appears to be protective [7] or, conversely, to be associated with higher persisting symptoms [7, 8]. So far, the underlying mechanisms of these symptoms remain unclear: the persistence of viral RNA and/or proteins has been demonstrated in some cases in Long COVID patients [10, 11, 12, 13, 14, 15; 16]. To date, it is not known whether this genetic or protein material persistence corresponds to a replicating virus or not. Such persistence or other potential mechanisms could lead to prolonged cytokine secretion, such as elevated levels of alpha and beta interferon [16], and to coagulation disorders [17], and tissue damage. Other research areas investigate hypotheses of a persistent inflammatory response including excessive mastocyte activation [18], of a defect of innate or adaptive immunity [19], of autoimmunity and of reactivation of latent viruses such as EBV [20]. Genetic and hormonal factors may be associated. The role of psychological factors that could lead to the continuation of symptoms is also discussed [21].

The Long COVID course fluctuates and usually lasts for months. Symptoms generally impact daily life functioning [22]. In comparison with hospitalised patients for whom data concerning the one year follow-up [23] are available (concerning the one year follow-up [23]), little is known about the long-term course of patients with prolonged COVID symptoms, the impact of the persistent symptoms on social, family and work life. Preliminary results obtained in a French E-cohort of Long COVID respondents to e-questionnaires, the COMPARE e-Cohort, suggest that less than 14% of monitored participants were in complete symptomatic remission after one year [24].

Our aim was, thus, to describe the 12-month evolution of Long COVID (LC) symptoms followed in an outpatient clinic in Paris and their impact on patients’ work lives and the factors associated with the relief of symptoms.

## Patients and Methods

### Setting

A post-acute COVID-19 clinic was established for outpatients complaining of persistent and/or recurrent COVID-19 symptoms since May 2020 in COCHIN HOTEL DIEU Hospital in Paris, affiliated with the Greater Paris Public Hospital (APHP). Several physicians took turns to receive patients. Patients were given a detailed medical assessment along with physical and biological examination. The following data were collected at baseline using a standardised questionnaire : age, sex, occupation, past medical history (including allergies, autoimmune disease, chronic disease, asthma, immunosuppression, lifestyle factors and exercise, anxiety and psychiatric history), body measurements, symptoms of acute COVID-19 onset, duration and treatment received, type, history and chronology of persistent and/or resurgent symptoms and their course (persistence, recurrence..), type of progression of persistent symptoms (unsustained or continuous), medical care and treatment, return to work and usual activities. A SARS-COV-2 serology was routinely requested. SARS-COV-2 RT-PCR assay on nasopharyngeal swabs and other bioassays were requested based on symptoms presented by participants. Additional tests and follow-up were provided to patients when needed.

### Ethics approval and consent to participate

Patients with the following inclusion criteria were asked to give a non-opposition be included in an observational study called PERSICOR: age> 18, an initial COVID-19 infection as defined by WHO COVID-19 Case definition [25]; at least two persistent or recurrent symptoms lasting for more than 2 months and occurring in the three months following initial COVID infection and absence of another obvious cause of symptoms, a definition corresponding to the post COVID-19 condition WHO clinical case definition [4]. Ethics approval for PERSICOR study was granted by the local Institutional Review Board of Henri-Mondor Hospital (Ethics Committee number 00011558, Approval number 2020-088).

### Definitions

Post COVID-19 condition was defined as described in the WHO clinical case definition of post COVID-19 condition [4] by: (1) a history of confirmed or probable SARS CoV-2 infection as described in the WHO COVID-19 Case definition [25], (2) symptoms occurring usually 3 months from the onset of COVID-19 lasting for at least 2 months, (3) including mainly fatigue, shortness of breath, cognitive dysfunction but also impact on everyday functioning (4) that cannot be explained by an alternative diagnosis.

#### Initial Long COVID-19 phase

was defined as the period between the first onset of Long COVID-19 symptoms after the initial COVID-19 episode and the first consultation at the outpatient Long COVID-19 clinic.

#### Strong evidence in favor of an initial SARS-CoV-2 infection

were defined as subjects having at the initial SARS infection either: (1) a positive PCR, (2) anosmia or ageusia, (3) typical COVID-19 lesions on chest CT scan, or (4) a close contact with a confirmed case.

### SARS-CoV-2 positive serology

Anti–SARS-CoV-2 spike IgG values were performed with commercially available tests and interpreted as positive according to the manufacturer’s recommendations. Only antibody measurements performed before any vaccination were considered. A post infection positive SARS-CoV-2 serology was defined by detecting one or more antibodies against SARS-CoV-2 S and/or N proteins at least once before any vaccination. A post infection negative SARS-CoV-2 serology was defined by the absence of detection antibodies against SARS-CoV-2 S and/or N proteins.

### Assessment at one year of initial COVID-19

One year after the initial long COVID-19 infection, participants included in the PERSICOR study were systematically contacted again and a standardised questionnaire was sent to them to evaluate at 12 months (+/- 1 month), the course of their symptoms, the diagnoses that were made, the treatments prescribed and their effect on the symptoms, the impact of the disease on patients’ social lives, work activities and family life. Patients were asked to rate their overall health on a score from 10 to 1 on a post-COVID-19 condition self-assessment scale that was designed by our team. Supplementary Materials A. explains how to use this score: 10 being the complete remission of symptoms, 8-9 having a significant improvement of symptoms with disappearance of most symptoms, 6-7 partial disappearance of symptoms, 3-5 persistence of symptoms and major limitation in daily life activities, 1-2 persistence of symptoms and dependence in daily life activities. Participants had to fill out this questionnaire and email it back or return it in a standard medical follow-up visit.

### Participants

To be included in this sub-analysis, participants were selected from the PERSICOR study based on the following criteria: having an initial confirmed or probable case of COVID-19 as defined by WHO COVID-19 case definition [25]; having their first COVID-19 episode between January 15, 2020 and May 21, 2021. Exclusion criteria were having only a suspected initial COVID-19 infection as defined by WHO COVID-19 case definition [25]; having developed a severe infection requiring intensive care support during the initial COVID-19 phase.

### Statistical analyses

Quantitative data were presented with mean, median, and interquartile ranges. For qualitative data, we used frequency distribution and relative frequency. 95% confidence intervals (CI) of proportions were calculated using the Clopper–Pearson interval by calculating quintiles from the beta distribution.

The symptoms observed were grouped into ten main classes: asthenia, neurocognitive and neurological, cardiothoracic, smell and taste, digestive, other ear-nose-throat symptoms, fevers or shivering, musculoskeletal, cutaneous-vascular and ophthalmic symptoms. The symptom coding of this study is detailed in Supplementary Materials B.

Cumulative percentage of participants reporting symptom remission (score at 10) was calculated using the reverse Kaplan–Meier method.

We then analysed the factors associated with a “significant improvement of symptoms or complete remission” at one year. A “significant improvement of symptoms or complete remission” was defined as giving an overall post COVID self-assessment score between 8 and 10, at 12 months. Variables studied were the participants’ baseline characteristics, type of symptoms at the initial COVID-19 episode, and type of symptoms at the first consultation for Long COVID (Supplementary Materials B), and SARS-CoV-2 serological result. We estimated the “prevalence rate ratios (PRR)” using univariate and adjusted PRRs multivariate general linear models (Poisson regression).

Multivariate analyses included the classes of symptoms at the first consultation for Long COVID, result of SARS-CoV-2 serology and were age- and sex-adjusted.

We performed a sensitivity analysis considering participants who had developed SARS-CoV-2 antibodies before vaccination or who had strong evidence in favor of SARS-CoV-2 infection, i.e. a positive PCR, anosmia or ageusia, typical COVID lesions on chest CT scan or contact with a confirmed case. Calculations were made with the software packages SAS (version 9.4) and R (version 4.2.1).

## Results

### Population characteristics

Among the 304 long COVID patients who reached one year follow-up at the date point, 296 could be contacted again, 231 (76%) answered a 12-month follow-up questionnaire. The patients’ characteristics are presented in Table 1.

**Table 1.** Characteristics of the patients and association with significant improvement or complete remission of symptoms at one year (univariate analysis)

| Background characteristics | Sample size | Significant improvement or complete remission at one year (score=8 to 10 on remission scale) | PRR of having significant improvement or complete remission |
| --- | --- | --- | --- |
|  | n (%) | n (%) |  |
|  | N=231 | N=86 |  |
| **Age at initial COVID-19 infection (%)** |  |  | **p=0.716** |
| under 40 | 65 (28.1%) | 23 (35.4%) | 1 |
| 40 and older | 166 (71.9%) | 63 (38.0%) | 1.07 (0.73-1.57) |
| **Sex (%)** |  |  | **p=0.836** |
| Female | 179 (77.5%) | 66 (36.9%) | 1 |
| Male | 52 (22.5%) | 20 (38.5%) | 1.04 (0.70-1.55) |
| **Body Mass Index (%)** |  |  | **p=0.927** |
| BMI<25 | 162 (70.1%) | 60 (37.0%) | 1 |
| BMI≥25 | 69 (29.9%) | 26 (37.7%) | 1.02 (0.71-1.47) |
| **Occupation (%)** |  |  | **p=0.838** |
| Health and social workers | 60 (26.0%) | 23 (38.3%) | 1 |
| Others | 171 (74.0%) | 63 (36.8%) | 0.96 (0.66-1.41) |
| **Tobacco consumption (%)** |  |  | **p=0.828** |
| No | 211 (91.3%) | 79 (37.4%) | 1 |
| Yes | 20 (8.7%) | 7 (35.0%) | 0.93 (0.51-1.73) |
| **Period of initial COVID-19 infection (%)** |  |  | **p=0.610** |
| Before June 2020 | 184 (79.7%) | 70 (38.0%) |  |
| After June 2020 | 47 (20.3%) | 16 (34.0%) | 0.89 (0.58-1.38) |
| **Atopy/Allergy/Asthma (%)** |  |  | **p=0.894** |
| No | 98 (42.4%) | 64 (36.4%) | 1 |
| Yes | 133 (57.6%) | 22 (40.0%) | 1.02 (0.73-1.44) |
| **Personal or familial autoimmune disease (%)** |  |  | **p=0.631** |
| No | 176 (76.2%) | 64 (36.4%) | 1 |
| Yes | 55 (23.8%) | 22 (40.0%) | 1.10 (0.75-1.62) |
| **Depression, anxiety history (%)** |  |  | **p=0.312** |
| No | 174 (75.3%) | 68 (39.1%) | 1 |
| Yes | 57 (24.7%) | 18 (31.6%) | 0.81 (0.53-1.22) |
| **Hospitalisation at initial COVID-19 infection (%)** |  |  | **p=0.067** |
| No | 178 (77.1%) | 72 (40.4%) | 1 |
| Yes | 53 (22.9%) | 14 (26.4%) | 0.65 (0.41-1.03) |
| **Number of symptoms at initial COVID-19 phase (%)** |  |  | **p=0.289** |
| Under 5 | 111 (48.1%) | 38 (34.2%) | 1 |
| 5 and over | 120 (51.9%) | 48 (40.0%) | 1.17 (0.83-1.64) |
| **SARS-CoV-2 positive serology at least once before vaccination (%)** |  |  | **p=0.005** |
| No | 85 (36.8%) | 22 (25.9%) | 1 |
| Yes | 146 (63.2%) | 64 (43.8%) | 1.67 (1.15-2.49) |
| CI: confidence interval |  |  |  |
| PRR: prevalence rate ratio obtained using Poisson regression |  |  |  |

Median age was 45 years (IQR: 38-53.), 77.5% were female. Median follow-up was 12.0 months (IQR: 10–14 months), and 63.2% had a positive SARS-CoV-2 serology, 26.0% were health workers, 57.6% had a history of atopy/allergy/asthma and 24.7% had a history of depression or anxiety prior to the long COVID-phase. At acute COVID-19 infection, 51.9% of participants presented at least five symptoms. Median follow-up after the acute episode of COVID-19 was 12.0 months (IQR: 11–13 months).

Within the subgroup of 85 participants lacking SARS-CoV-2-antibodies or who had not performed SARS-COV-2 serology, 71.8% (61/85) had strong evidence in favor of an initial SARS infection: 19 had a positive PCR, 48 reported anosmia or ageusia, 13 had typical COVID-19 lesions on chest CT scan and 19 had been in close contact with a confirmed case.

### Course of symptoms over time

The prevalence rates of symptoms are presented in Table 2 and Figure 1 at three different time points: acute COVID-19 episode, L ong COVID phase and one year follow-up.

**Table 2.** Evolution of the prevalence of symptoms at the three phases of the study: initial COVID-19 episode, initial Long COVID-19 phase, one year follow-up

| Symptoms | Initial COVID-19 episode |  | Initial long COVID -19 phase |  | One year follow-up |  |
| --- | --- | --- | --- | --- | --- | --- |
|  | N | % 95%CI | N | % 95%CI | N | % 95%CI |
| Smell and taste | 144 | 62.3%(55.7%-68.6%) | 82 | 35.5%(29.3%-42.0%) | 92 | 39.8%(33.5%-46.5%) |
| Other ear-nose-throat disorders | 183 | 79.2%(73.4%-84.3%) | 134 | 58.0%(51.4%-64.5%) | 146 | 63.2%(56.6%-69.4%) |
| Asthenia | 155 | 67.1%(60.6%-73.1%) | 178 | 77.1%(71.1%-82.3%) | 192 | 83.1%(77.7%-87.7%) |
| Fevers or Shivering | 196 | 84.8%(79.6%-89.2%) | 37 | 16.9% (12.3%-22.3%) | 75 | 32.5%(26.5%-38.9%) |
| Cardiothoracic | 193 | 83.5%(78.1%-88.1%) | 187 | 81.0%(75.3%-85.8%) | 180 | 77.9%(72.0%-83.1%) |
| Neurological and neurocognitive | 169 | 73.2%(67.0%-78.8%) | 193 | 83.5%(78.1%-88.1%) | 217 | 93.9%(90.0%-96.6%) |
| Digestive | 86 | 37.2%(31.0%-43.8%) | 93 | 40.3%(33.9%-46.9%) | 124 | 53.7% (47.0%-60.2%) |
| Musculoskeletal | 86 | 37.2%(31.0%-43.8%) | 93 | 40.3%(33.9%-46.9%) | 182 | 78.8%(72.9%-83.9%) |
| Cutaneous and vascular | 29 | 12.6%(8.6%-17.5%) | 72 | 31.2%(25.3%-37.6%) | 142 | 61.5%(54.9%-67.8%) |
| Ophthalmic | 26 | 11.3%(7.5%-16.1%) | 48 | 20.8%(15.7%-26.6%) | 103 | 44.6%(38.1%-51.2%) |

*Other ear-nose-throat: tinnitus, vestibular involvement, odynophagia, rhinorrhea, dysphagia or other otorhinolaryngologic symptoms*
*Cardiothoracic: dyspnea, cough, thoracic, chest tightness, tachycardia, bradycardia or desaturation*
*Other neurological disorders: difficulty swallowing, difficulty articulating, urinary retention, thermoregulation disorders, deafness, dysphonia, thermoregulation, insomnia/hypersomnia*
*Psychological disorders: anxiety, emotionality, thymic disorders, mood disorders*
*Digestive: diarrhea, abdominal pains, constipation, gastroparesis or vomiting/nausea*
*Ophthalmic: dry eyes, cloudy vision, conjunctivitis, orbit pain or blurred vision*

**Figure 1:**
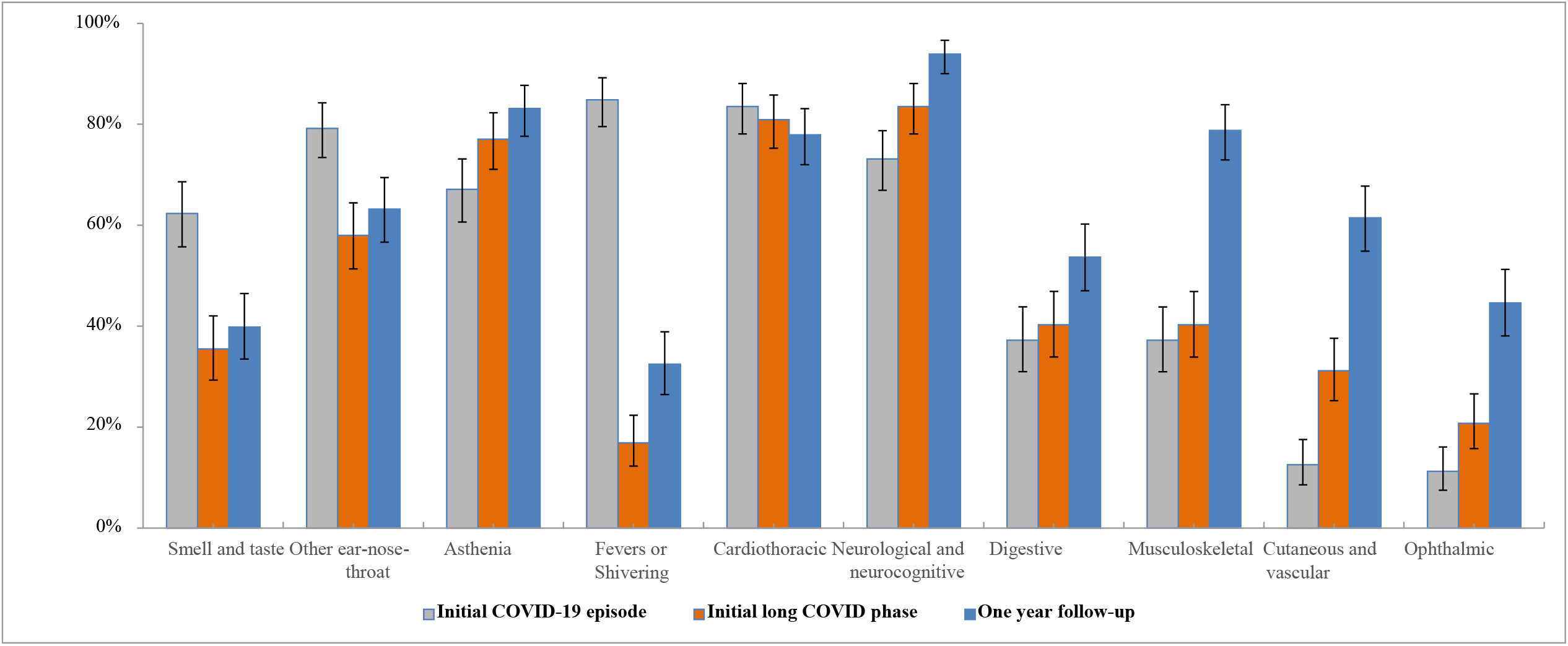
Dynamics of course of the three phases of the study: initial COVID-19 infection, initial Long COVID phase and one year follow-up (Proportion and 95% confidence interval) **Smell and taste symptoms:** Ageusia, anosmia **Other ear-nose-throat:** Tinnitus, vestibular involvement, odynophagia, rhinorrhoea, dysphagia or other otorhinolaryngologic symptoms **Cardiothoracic**: Dyspnea, cough, thoracic pain, chest tightness, tachycardia, bradycardia or desaturation **Neurological and neurocognitive**: Neurocognitive, sensory disorders, other neurological **Neurocognitive**: Concentration and attention disorders (bradypsychia), immediate memory disorders, mood disorders, insomnia/hypersomnia **Sensory disorders**: Tingling, burning, neurogenic pain or balance disorders or paresthesia **Other neurological**: Difficulty swallowing, difficulty articulating, bladder retention, thermoregulation disorders, deafness, dysphonia, or thymic disorders Musculoskeletal : Myalgias, muscle weakness, arthralgias or tenosynovitis **Cutaneous and vascular**: Urticaria, eczema, subcutaneous hematoma or vascular inflammation **Ophthalmic**: Dry eyes, cloudy vision, conjunctivitis, orbit pain or visual blurring

### Course of symptoms between the acute COVID-19 episode and first L ong COVID phase

Between acute COVID-19 episode and initial Long COVID phase, the prevalence rate of symptoms decreased for smell and taste (62.3% to 35.5%), for fevers/shivering (84.8% to 18.2%) and for other ENT (ear - nose - throat) symptoms (79.2% to 58.0%). Conversely, the prevalence rate increased for neurocognitive and neurological symptoms (73.2% to 83.5%), and for cutaneous and vascular symptoms (12.6% to 31.2%). The prevalence rate remained stable until 12 months for skeletal muscle, digestive and cardiothoracic symptoms, and asthenia (Table 2).

### Course of symptoms between initial long COVID phase and 12 month follow-up

Figure 2 and Supplementary Materials C highlight symptom-specific courses between initial Long COVID phase and one year follow-up. Between the initial Long COVID phase and the 12-month follow-up, the symptoms persisted and even increased, for asthenia (77.1% to 83.1%), neurocognitive and neurological symptoms (83.5% to 93.9%), cardiothoracic (81.0% to 77.9%), musculoskeletal (40.3% to 78.8%), cutaneous and vascular (31.2% to 61.5%) and ophthalmic symptoms (20.8% to 44.6%). Among other ENT symptoms, tinnitus or vestibular involvement increased (16.0% to 42.9%). Likewise, psychological symptoms increased (18.7% to 68.4%). However, regarding cardiothoracic symptoms, thoracic pain, oppression and tachycardia decreased (67.1% to 50.6%).

**Figure 2:**
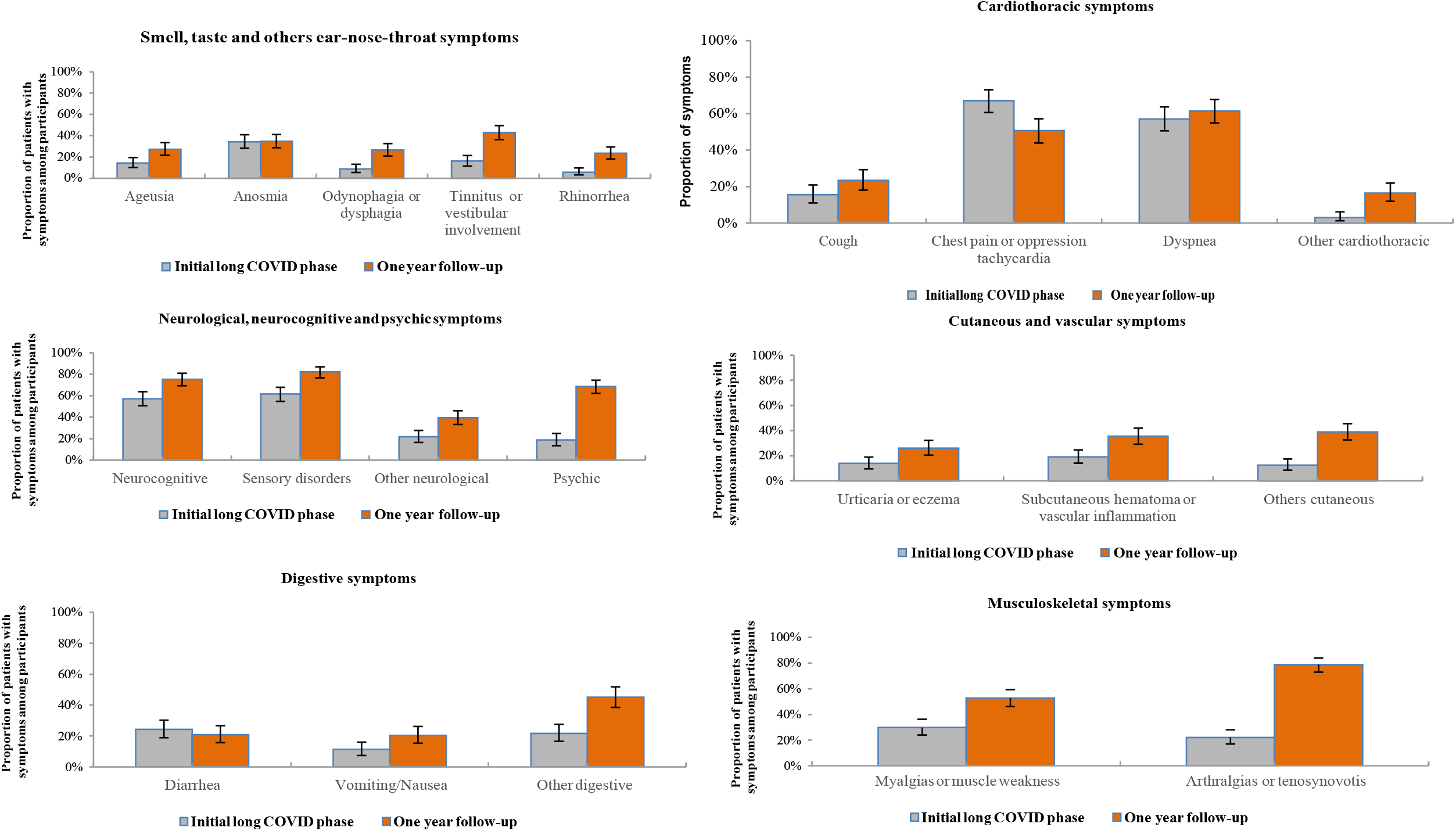
Evolution by symptom class: initial long COVID phase and one year follow up (Proportion and 95%confidence interval) **Others cardiothoracic**: Hypertension, orthostatic hypotension, asthma **Others** **digestives**: Constipation, flatulence-bloat, gastroesophageal reflux **Others cutaneous:** Peeling skin, skin rash, hair loss, skin dryness **Others neurological**: Difficulty swallowing, difficulty articulating, bladder retention thermoregulation disorders, deafness, dysphonia, thermoregulation

### Changing health conditions, remission rate at 12 months, care and impact on daily life

Figure 3 highlights changing health conditions at 12 months, as evaluated by the participants and as compared to their pre COVID-19 status along with the scale used. At 12-month follow-up, the mean of post-COVID-19 condition self-assessment score (median; interquartile range) was 6.6 (7.0; 6.0 to 8.0). Supplementary D presents the cumulative percentages of participants reporting complete symptom remission (score at 10). The cumulative percentages of complete remission at 12 months were 8.7%. A percentage of 28.6% felt a significant improvement of their symptoms (score at 8-9) while 62.7% had a minor or no significant improvement of symptoms (score <8).

**Figure 3.**
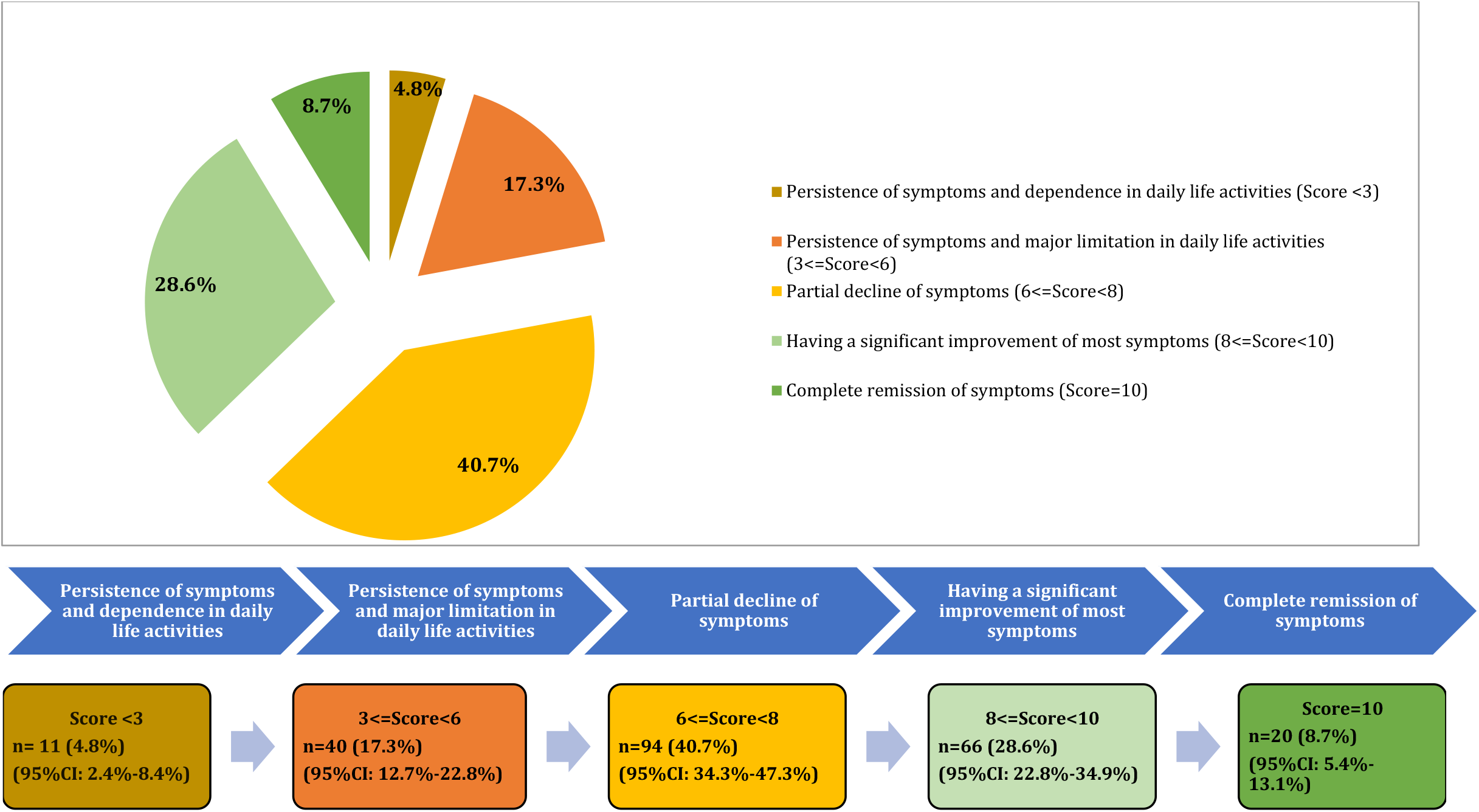
Perception of overall health condition in participants at one year follow-up. Supplementary Materials A gives instructions to use this score

Another 40.7% had a partial improvement of their persistent symptoms (score at 6-<8). In total, 17.3% reported a persistence of most symptoms and a major limitation in daily life activities (3-<6), 4.8% had a persistence of all the symptoms and a dependence in daily life activities (1-<3). Table 3 reports the impact of Long COVID on daily life. Before the acute phase of COVID-19, 193/231 (83.5%) had a professional activity, 14/231 (6.1%) were retired and 24/231 (10.4%) were unemployed.

**Table 3.** Impact of long-term persistent symptoms on professional and social life, and on daily activities at one year

| Activities | Sample size | Percentage (IC 95%) |
| --- | --- | --- |
| **Professional activities** |  |  |
| **Professional activities before acute Covid-19 (n=231)** |  |  |
| No | 38 | 16.5% (11.9% - 21.9%) |
| Yes | 193 | 83.5% (78.1% - 88.1%) |
| **Work interruption during persistent Covid-19 symptoms (n=193) \*** |  |  |
| No | 73 | 37.8% (31.0% - 45.1%) |
| Yes | 120 | 62.2% (54.9% - 69.0%) |
| **Returning to work after stopping (n=120) \*\*** |  |  |
| No | 40 | 33.3% (25.0% - 42.5%) |
| Yes | 80 | 66.7% (57.5% - 75.0%) |
| **Professional activities at one-year follow-up (n=231)** |  |  |
| No | 77 | 33.3% (27.3% - 39.8%) |
| Yes | 154 | 66.7% (60.2% - 72.7%) |
| **Working part-time at one year (n=154\*\*\*)** |  |  |
| Part-time | 104 | 67.5% (59.5% - 74.8%) |
| Full-time | 50 | 32.5% (25.2% - 40.5%) |
| **Domestic and social activities** |  |  |
| **Resumption of domestic activities at one year (n=231)** |  |  |
| No | 58 | 25.1% (19.7% - 31.2%) |
| Part-time | 116 | 50.2% (43.6% - 56.8%) |
| Full-time | 57 | 24.7% (19.3% - 30.8%) |
| **Sports activities at one year (n=231)** |  |  |
| Unable to exercise | 57 | 24.7% (19.3% - 30.8%) |
| Difficult to exercise | 98 | 42.4% (36.0% - 49.1%) |
| Possible to do sports | 67 | 29.0% (23.2% - 35.3%) |
| Did not try to exercise | 9 | 3.9% (1.8% - 7.3%) |
| **Ability to drive a car at one year (n=231)** |  |  |
| Unable to do it | 57 | 24.7% (19.3% - 30.8%) |
| Difficult to do it | 98 | 42.4% (36.0% - 49.1%) |
| Possible to do it | 67 | 29.0% (23.2% - 35.3%) |
| Did not try to do it | 9 | 3.9% (1.8% - 7.3%) |
| **Ability to read at one year (n=231)** |  |  |
| Unable to do it | 8 | 3.5% (1.5% - 6.7%) |
| Difficult to do it | 74 | 32.0% (26.1% - 38.5%) |
| Possible to do it | 138 | 59.7% (53.1% - 66.1%) |
| Did not try to do it | 11 | 4.8% (2.4% - 8.4%) |
| *Stopping work among participants in activities before Covid -19 |  |  |
| **Returning to work among participants who stopped working during long COVID phase |  |  |
| *** Rate of return to work among participants who were employed before Covid -19 |  |  |

Among the 193 participants who were working before COVID-19 infection, 120 respondents (62.2%) had to stop working at least once during the Long COVID phase. Among those who stopped work, 154/231 (66.7%) resumed their professional activities at one year; 154/231 (66.7%) had a professional activity, versus 193/231 (83.5%) before the acute phase of COVID. Among those who held a professional activity at one year, less than a third were working full time.

Otherwise, less than a quarter, 57/231 (24.7%), resumed routine domestic activities after 12 months. Among those who tried to do a sport activity, only 67/222 (30.2%) could do it without any difficulty. Among those who tried to drive a car, 169/206 (72.3%) could do it without any difficulty, and among those who tried to read, 138/220 (62.7%) could do it without any difficulty.

### Factors associated with “significant improvement of symptoms”

At the 12 month assessment, 37.2% (95%CI: 31.0%-43.8%) of the participants overall noted a significant improvement of their general health condition or a complete remission on the remission scale. Table 1 and Figures 4 and 5 present factors associated with a significant improvement of symptoms (score 8 to 10) in univariate and multivariate analysis.

**Figure 4:**
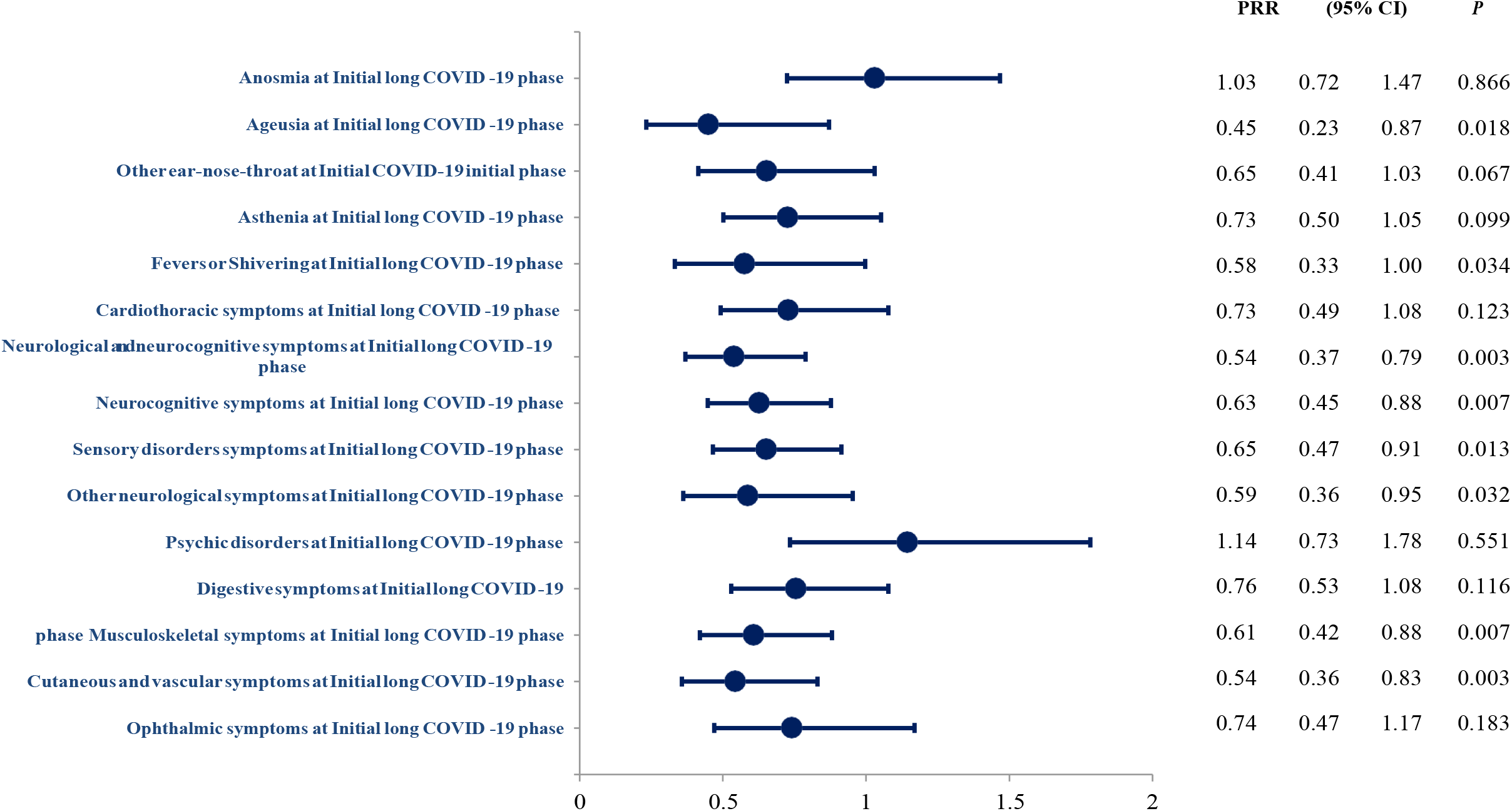
Symptom persistence at COVID-19 initial phase associated with significant improvement of symptoms or complete remission (Univariate analysis) **PRR** = prevalence rate ratio obtained using Poisson regression **Other ear-nose-throat:** Tinnitus, vestibular involvement, odynophagia, rhinorrhoea, dysphagia or others otorhinolaryngologic symptoms **Cardiothoracic:** Dyspnea, cough, thoracic pain, chest tightness, tachycardia, bradycardia or desaturation **Neurological and neurocognitive**: Neurocognitive, sensory disorders, others neurological **Neurocognitive**:Concentration and attention disorders (bradypsychia), immediate memory disorders **Sensory disorders**: Tingling, burning, neurogenic pain or balance disorders or paraesthesia **Others neurological**: Difficulty swallowing, difficulty articulating, bladder retention thermoregulation disorders, deafness, dysphonia,, insomnia/hypersomnia **Psychic disorders**: Anxiety, emotionality, thymic disorders, mood disorders **Digestive**: Diarrhea, abdominal pains, constipation, gastroparesis or Vomiting/Nausea **Musculoskeletal**: Myalgias, muscle weakness, arthralgias or tenosynovitis **Cutaneous and vascular**: Urticaria, eczema, subcutaneous hematoma or vascular inflammation **Ophthalmic**: Dry eyes, cloudy vision conjunctivitis, orbit pain or visual blurring

**Figure 5:**
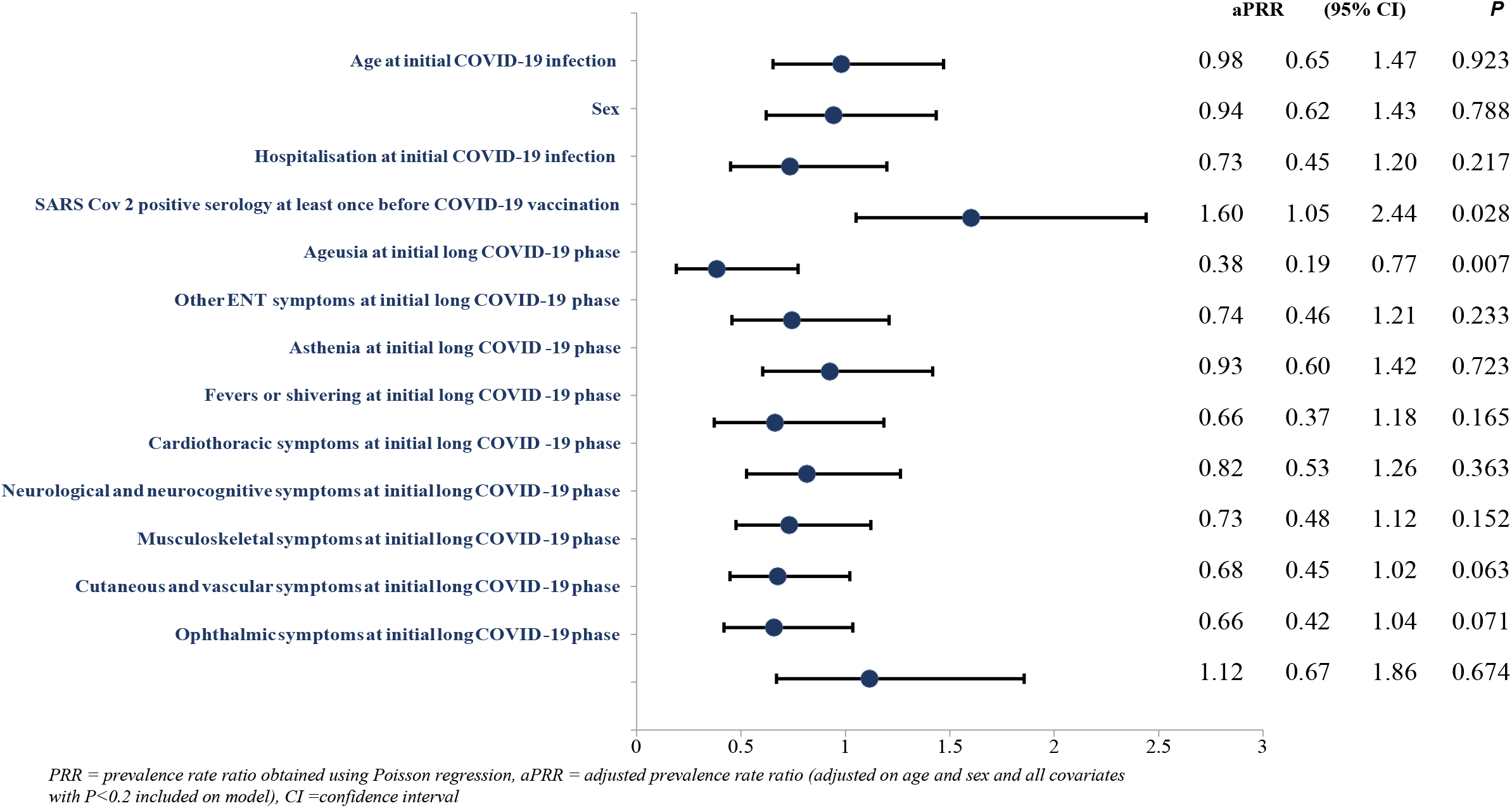
Factors associated with significant improvement of symptoms or complete remission (multivariate analysis) **Other ear-nose-throat:** Tinnitus, vestibular involvement, odynophagia, rhinorrhea, dysphagia or others ENT symptoms **Cardiothoracic:** Dyspnea, cough, thoracic pain, chest tightness, tachycardia, bradycardia or desaturation **Neurological and neurocognitive**: Neurocognitive, sensory disorders, other neurological **Neurocognitive:**Concentration and attention disorders (bradypsychia), immediate memory disorders **Sensory disorders:** Tingling, burning, neurogenic pain or balance disorders or paresthesia **Others neurological**: Difficulty swallowing, difficulty articulating, urinary retention thermoregulation disorders, deafness, dysphonia, insomnia/hypersomnia **Musculoskeletal**: Myalgias, muscle weakness, arthralgias or tenosynovitis **Cutaneous and vascular**: Urticaria, eczema, subcutaneous hematoma or vascular inflammation **Ophthalmic**: Dry eyes, cloudy vision conjunctivitis, orbit pain or visual blurring

#### *<u>In univariate analysis</u>* (Figure 4)

Participants who had SARS-CoV-2 antibodies at least once had an increased probability of significant improvement at 12-month follow-up as compared to those with a negative serology (PRR: 1.67, p=0.005). Participants who presented at the initial Long COVID phase with ageusia (PRR: 0.45, p=0.008), neurocognitive symptoms (PRR: 0.54, p=0.003), musculoskeletal symptoms (PRR: 0.61, p=0.007), and cutaneous and vascular symptoms (PRR: 0.48, p=0.003) had a decreased probability of significant improvement at one year.

Among those who had strong clinical arguments documenting the initial SARS-CoV-2 infection, ageusia (PRR: 0.41, p=0.007), neurocognitive (PRR: 0.55, p=0.002) and musculoskeletal (PRR: 0.60, p=0.006) symptoms at initial Long COVID phase had a decreased probability of significant improvement at one year. Participants who had SARS-CoV-2 antibodies at least once had an increased probability of significant improvement at 12-month follow-up as compared to those with a negative serology (PRR: 1.41, p=0.092).

#### *<u>In multivariate analysis</u>* (Figure 5)

In adjusted analysis, participants who had SARS-CoV-2 antibodies at least once before COVID-19 vaccination had a higher probability of significant improvement of symptoms at one year than those who had no SARS-CoV-2 antibodies at any consultation (aPRR: 1.60, 95%CI: 1.05–2.44, P = 0.028).

Participants who had ageusia at the initial Long COVID phase (aPRR: 0.38; 95%CI: 0.19-0.77, p=0.007) had a lower probability of significant improvement of symptoms at one year than those who did not develop ageusia during the Long COVID phase.

Among participants who had strong evidence in favor of initial SARS-CoV-2 infection, ageusia at initial Long COVID phase was associated with decreased likelihood of significant improvement at one year (aPRR: 0.37 P=0.004).

## Discussion

Our study, that focused on the 12 month follow-up of 231 long COVID patients, showed that only 37.2% of the participants rated a significant improvement of their symptoms or felt in complete remission at one year. In the majority of the patients, some symptoms such as asthenia and neurocognitive symptoms remained highly prevalent at one year. Only 77.2% of the patients who worked before COVID returned to work at one year and, among these, 32.5% worked part-time. Interestingly, the factors associated with persistence of symptoms were the following: not having developed SARS-CoV-2 antibodies, and having persistent ageusia over the Long COVID phase. Concerning the persistence of symptoms at one year, our results are concordant with the few published studies, namely the Compare e-cohort study showing that at one year, 89% of the patients with Long COVID still complained of persisting symptoms [24]. In a recent meta-analysis of 19 studies, encompassing a total of 11,324 patients, approximately 30% of patients reported long-term persistence of symptoms after COVID-19, among which neurocognitive, psychiatric and sleep disorders were the most frequent. This is particularly preoccupying for the future [26].

However, no study has yet described the clinical and biological factors linked to symptom remission in patients with a diagnosis of Long COVID. What’s more, the impact of these persistent symptoms on resuming work and everyday activities has rarely been described.

Having developed SARS-CoV-2 antibodies seems linked to greater symptom remission. All the patients in our study met the WHO definitions for both initial and Long COVID. Among them, 63% had a positive SARS-CoV-2 serology before any vaccination, while 34% never had detectable antibodies and 3% had no serological tests performed.

The adaptive immune system plays a key role in controlling most viral infections and among the three fundamental components of the adaptive immune system (B cells, CD4+ T cells, and CD8+ T cells), humoral responses induced by B Cells are particularly important to fight viral infections. We found that the probability of having a significant improvement at one year was higher in those who had SARS COV2 antibodies than in those who did not (43.8% vs 25.9%). In other words, the capacity of having developed SARS-CoV-2 antibodies was linked to a better outcome. This suggests that one of the mechanisms to explain Long COVID occurrence might be the inability to mount an efficacious adaptive immune response. This point is debated in the scientific community. An early study showed that a risk factor for the persistence of symptoms after an initial COVID infection was low SARS-CoV-2 antibodies level [7]. This finding was expanded on by another pilot study showing that half of the patients seeking medical help for post-acute COVID-19 syndrome lack a humoral response against SARS-CoV-2, including some PCR+ patients. However, in the latter study, some patients did not have virologically documented infections [19].

Another study has identified that in patients with Long COVID, as compared to patients with COVID resolved without sequelae, N-specific CD8+ T cell responses were lower and waning more rapidly at month 4 [27]. However, this study showed that there was no difference in antibody neutralisation titers between both groups.

Naturally, we may suppose that patients without SARS-CoV-2 antibodies suffered from another disease than COVID-19 [21]. To prevent such possibility, we excluded patients who only had a suspected diagnosis of COVID and included only those with a documented or probable COVID diagnosis as defined by WHO and excluded also those who had an alternative diagnosis to COVID.

Moreover, among the subgroup lacking SARS-CoV-2 antibodies, we found that 71.8 % had strong evidence in favor of the initial SARS infection (either a positive PCR, anosmia or ageusia; or typical lesions on chest CT scan; or close contact with a confirmed case). Overall, 89.6% of our population either developed SARS-CoV-2 antibodies before vaccination or had strong evidence in favor of initial SARS infection. The sensitivity analysis performed on patients who developed SARS-CoV-2 antibodies before vaccination or patients who had a strong argument to document a SARS-CoV-2 infection showed that the factors associated with significant improvement or complete symptom remission at one year were the same as the factors observed in the analysis on overall participants.

The absence of a humoral response after SARS-COV-2 is not an isolated phenomenon. Several studies have shown that following a minor infection, mostly in children, a certain percentage of subjects did not develop antibodies or lost them quickly [28].

This finding reinforces the hypothesis that one of the mechanisms of PACS could be an inability by the host to clear the virus and viral persistence in reservoirs.

Additionally, we found that persistent ageusia in the Long COVID phase (with or without smell disturbance) was associated with the absence of symptom remission at one year.

This finding cannot be easily explained and requires looking into the pathophysiology of ageusia in COVID-19. A review article has compiled 47 articles discussing the hypothetical mechanisms of action and etiopathogenesis of ageusia in COVID-19 patients [29]. Regarding transient ageusia occurring during acute COVID, the most likely causes seem to be a direct viral neural invasion of the olfactory and gustatory nerves, viral cytotoxicity to taste buds, angiotensin II imbalance, augmented pro-inflammatory cytokines, and disturbances in salivary glands and sialic acid [30]. However, chronic ageusia, especially when combined with other neurological symptoms, seems to be caused by damage to one or more cranial nerves, to the brainstem or to the cerebral cortex. The taste nervous system is based on three pairs of cranial nerves: the facial nerve, the glosso-pharyngeal nerve and the vagus nerve. The nerve fibers carried by these three nerves all end in the nucleus of the solitary tract, then the projections towards the thalamus and the insula are made [30].

Since 2021, 18 FDG brain PET 18F-fluorodeoxyglucose (FDG)-positron emission tomography (PET-CT scan) studies have shown that the brainstem constitutes one of the areas most affected by neuroinflammation during Long COVID [31, 32]. PET-CT scans usually reveal a profile of brain hypometabolism in Long COVID patients reporting neurocognitive impairment with specific areas affected in the brain stem such as the rectal/orbital gyrus, the temporal lobes, including the amygdala and the hippocampus, the thalamus, the pons/medulla brainstem, the insula, and the bilateral cerebellum. The right regions of the brain stem seem more frequently affected than the left zones. One of these studies has shown that a significant hypometabolism in the parahippocampal gyri and orbitofrontal cortex hypometabolism characterised patients with persistent ageusia and anosmia [32].

Hence, ageusia might be a proxy for brainstem involvement and patients with brainstem involvement might have a less reversible form of Long COVID. We did not find any direct correlation between ageusia and neurocognitive disorders, which may be explained by the very high prevalence of neurocognitive disorders within our population.

Since our study included a majority of female patients as in all Long COVID cohorts up to now, we did not find that the female sex was associated with a poor long-term outcome, as previously reported in the long-term follow-up of hospitalised patients [8, 9].

### Limitations

Significant limitations can be found in our study. First, it was a monocentric study performed in a centre of care for Long COVID patients. Hence, our study might not be representative of the whole population of Long COVID patients, which probably includes less severe patients suffering from a few symptoms only, such as isolated anosmia or fatigue.

Second, we conducted a real-life study for which cases of COVID-19 were not all virologically proven. SARS CoV-2 serology was performed in the standard of care at different times compared to the initial COVID-19 episode, using different methods. As a rule, serology testing was requested when the patient first came to the treatment centre but when a serology test was already available in the patient’s record, it was not repeated. It is worth noting that the time taken to complete serology testing from the initial COVID episode did not differ whether the patients had a positive or a negative serology test (median of 2 months in both groups).

### Strengths

One of the strengths of the current study lies in the fact that we achieved a high level of responses from our patients to the one-year questionnaire, which gives us a sufficient sample to study the factors associated with prognosis. Additionally, our study constitutes one of the few studies, assessing, to date, the return to work and daily activities at one year.

In conclusion, in Long COVID patients consulting at a Long COVID support centre, the one year follow-up shows that less than 10% of the patients are in complete remission of their symptoms while the majority remained disabled in their daily activities, mainly because of fatigue and neurocognitive disorders. Our data show that having developed SARS COV-2 antibodies independently of vaccination seems associated with a more favorable course, which could lead to specific immunotherapeutic interventions. Soon, we hope to be able to describe the course of the disease at 3 years and that most of our patients will enjoy a favorable outcome of their disease.

## Data Availability

All data produced in the present study are available upon reasonable request to the authors

## Acknowledgments

We would like to thank the patient associations and patients who donated to the association ESPOIRS which sponsored and funded this study.

