## Supplementary materials for "Factors associated with release relief of Long COVID symptoms at 12-Months and their impact on daily life"

**Supplementary Materials A. Patient post-COVID-19 condition self-assessment scale**

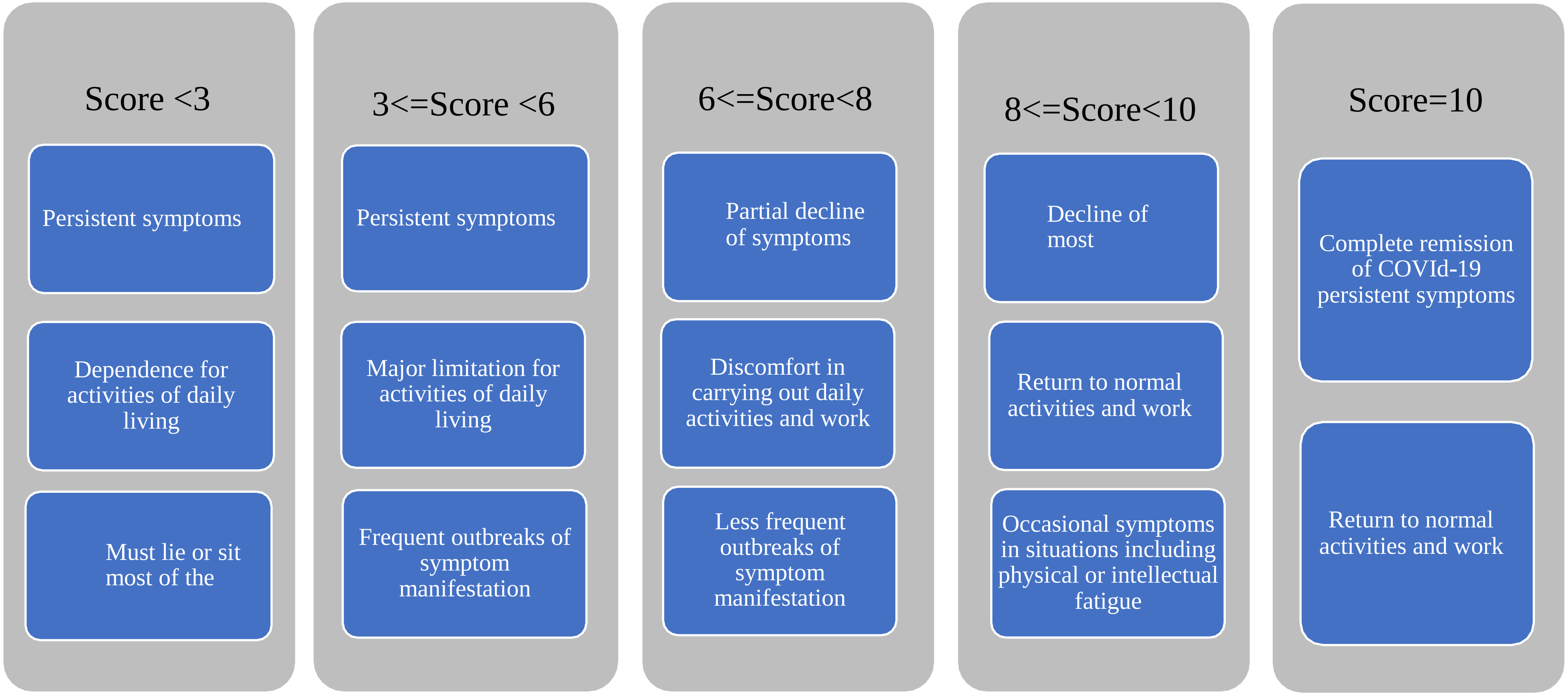

Supplementary Materials B. Study symptom coding

| Study symptom coding | Definition of symptoms group |
|---|---|
| **Smell and taste symptoms** | Aguesia, anosmia |
| **Other ear-nose-throat symptoms** | Tinnitus, vestibular involvement, odynophagia,rhinorrhea, dysphagia or other o torhinolaryngologic symptoms |
| **Asthenia** | Asthenia or fatigue |
| **Fevers or Shivering** | Fevers or shivering |
| **Cardiothoracic symptoms** | Dyspnea, cough, thoracic pain and oppression, tachycardia, bradycardia or desaturation |
| **Neurological and neurocognitive symptoms** | |
| ***Neurocognitive*** | Concentration and attention disorders (bradypsychia), immediate memory disorders, insomnia/hypersomnia |
| ***Sensory disorders*** | Tingling, burning, neurogenic pain or balance disorders or paresthesia |
| ***Other neurological*** | Difficulty swallowing, difficulty articulating, urinary retention, thermoregulation disorders, deafness, dysphonia, thermoregulation |
| **Psychic disorders** | Anxiety, emotionality, thymic disorders, mood disorders |
| **Digestive symptoms** | Diarrhea, abdominal pain, constipation, gastroparesis or vomiting/nausea |
| **Musculoskeletal** | Myalgias, muscle weakness, arthralgias or tenosynovitis |
| **Cutaneous and vascular symptoms** | Urticaria, eczema, subcutaneous hematoma or vascular inflammation |
| **Ophthalmic symptoms** | Dry eyes, cloudy vision, conjunctivitis, orbit pain or visual blurring |

Supplementary Materials C: Detail of symptoms by class between initial long COVID phase ant one year follow-up

| | Initial long COVID -19 phase | | | | | One year follow-up | | | | |
|---|---|---|---|---|---|---|---|---|---|---|
| Symptoms | N | % | 95%CI | | | N | % | 95%CI | | |
| Smell and taste | 82 | 35.5% | 29.3% | - | 42.0% | 92 | 39.8% | 33.5% | - | 46.5% |
| Ageusia | 33 | 14.3% | 10.0% | - | 19.5% | 63 | 27.3% | 21.6% | - | 33.5% |
| Anosmia | 79 | 34.2% | 28.1% | - | 40.7% | 80 | 34.6% | 28.5% | - | 41.2% |
| Other ear-nose-throat | 53 | 22.9% | 17.7% | - | 28.9% | 146 | 63.2% | 56.6% | - | 69.4% |
| Odynophagia or dysphagia | 20 | 8.7% | 5.4% | - | 13.1% | 61 | 26.4% | 20.8% | - | 32.6% |
| Tinnitus or vestibular involvement | 37 | 16.0% | 11.5% | - | 21.4% | 99 | 42.9% | 36.4% | - | 49.5% |
| Rhinorrhea | 13 | 5.6% | 3.0% | - | 9.4% | 54 | 23.4% | 18.1% | - | 29.4% |
| Asthenia | 178 | 77.1% | 71.1% | - | 82.3% | 192 | 83.1% | 77.7% | - | 87.7% |
| Fevers or Shivering | 39 | 16.9% | 12.3% | - | 22.3% | 75 | 32.5% | 26.5% | - | 38.9% |
| Cardiothoracic | 187 | 81.0% | 75.3% | - | 85.8% | 180 | 77.9% | 72.0% | - | 83.1% |
| Cough | 36 | 15.6% | 11.2% | - | 20.9% | 54 | 23.4% | 18.1% | - | 29.4% |
| Thoracic pain, chest thightness and tachycardia | 155 | 67.1% | 60.6% | - | 73.1% | 117 | 50.6% | 44.0% | - | 57.3% |
| Dyspnea | 132 | 57.1% | 50.5% | - | 63.6% | 142 | 61.5% | 54.9% | - | 67.8% |
| Other cardiothoracic | 7 | 3.0% | 1.2% | - | 6.1% | 38 | 16.5% | 11.9% | - | 21.9% |
| Neurological and neurocognitive | 193 | 83.5% | 78.1% | - | 88.1% | 212 | 91.8% | 87.5% | - | 95.0% |
| Neurocognitive | 132 | 57.1% | 50.5% | - | 63.6% | 174 | 75.3% | 69.2% | - | 80.7% |
| Sensory disorders | 142 | 61.5% | 54.9% | - | 67.8% | 190 | 82.3% | 76.7% | - | 87.0% |
| Other neurological | 50 | 21.6% | 16.5% | - | 27.5% | 91 | 39.4% | 33.0% | - | 46.0% |
| Psychic | 36 | 18.7% | 13.4% | - | 24.9% | 158 | 68.4% | 62.0% | - | 74.3% |
| Digestive | 93 | 40.3% | 33.9% | - | 46.9% | 124 | 53.7% | 47.0% | - | 60.2% |
| Diarrhea | 56 | 24.2% | 18.9% | - | 30.3% | 48 | 20.8% | 15.7% | - | 26.6% |
| Vomiting/Nausea | 26 | 11.3% | 7.5% | - | 16.1% | 47 | 20.3% | 15.3% | - | 26.1% |
| Other digestive | 50 | 21.6% | 16.5% | - | 27.5% | 104 | 45.0% | 38.5% | - | 51.7% |
| Musculoskeletal | 93 | 40.3% | 33.9% | - | 46.9% | 182 | 78.8% | 72.9% | - | 83.9% |
| Myalgias or muscle weakness | 69 | 29.9% | 24.0% | - | 36.2% | 122 | 52.8% | 46.2% | - | 59.4% |
| Arthralgias or tenosynovitis | 51 | 22.1% | 16.9% | - | 28.0% | 182 | 78.8% | 72.9% | - | 83.9% |
| Cutaneous and vascular | 72 | 31.2% | 25.3% | - | 37.6% | 142 | 61.5% | 54.9% | - | 67.8% |
| Urticaria or eczema | 32 | 13.9% | 9.7% | - | 19.0% | 60 | 26.0% | 20.4% | - | 32.1% |
| Subcutaneous hematoma or vascular inflammation | 44 | 19.0% | 14.2% | - | 24.7% | 82 | 35.5% | 29.3% | - | 42.0% |
| Other cutaneous | 29 | 12.6% | 8.6% | - | 17.5% | 90 | 39.0% | 32.6% | - | 45.6% |
| Ophthalmic | 48 | 20.8% | 15.7% | - | 26.6% | 103 | 44.6% | 38.1% | - | 51.2% |

Supplementary D: Cumulative percentage of participants reporting symptom remission at one year

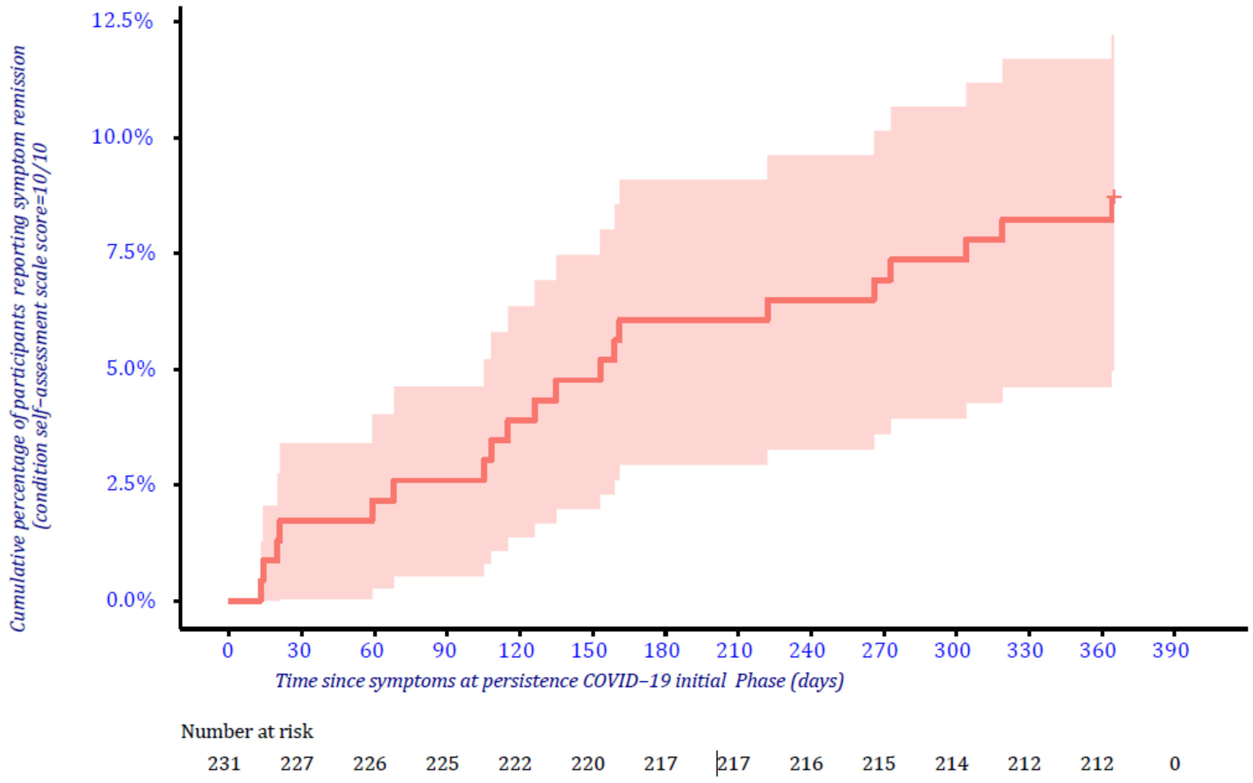
